# Evaluation of a microRNA-based Risk Classifier Predicting Cancer-Specific Survival in Renal Cell Carcinoma with Tumor Thrombus of the Inferior Vena Cava

**DOI:** 10.1101/2022.09.03.22279559

**Authors:** Mischa J. Kotlyar, Markus Krebs, Maximilian Burger, Hubert Kübler, Ralf Bargou, Susanne Kneitz, Wolfgang Otto, Johannes Breyer, Daniel C. Vergho, Burkhard Kneitz, Charis Kalogirou

## Abstract

**Background:** Clear cell renal cell carcinoma extending into the inferior vena cava (ccRCC^IVC^) represents a clinical high-risk setting. However, there is substantial heterogeneity within this patient subgroup regarding survival outcomes. Previously, members of our group developed a microRNA(miR)-based risk classifier – containing miR-21, miR-126 and miR-221 expression – which significantly predicted cancer-specific survival (CSS) of ccRCC^IVC^ patients.

**Methods:** Examining a single-center cohort of tumor tissue from n = 56 patients with ccRCC^IVC^, we measured expression levels of miR-21, miR-126 and miR-221 by qRT-PCR. Prognostic impact of clinicopathological parameters and miR expression were investigated via univariate and multivariate cox regression. Referring to the previously established risk classifier, we performed Kaplan Meier analyses for single miR expression levels and the combined risk classifier. Cut-off values and weights within the risk classifier were taken from the previous study.

**Results:** miR-21 and miR-126 expression were significantly associated with lymphonodal status at time of surgery, development of metastasis during follow-up, and cancer-related death. In Kaplan Meier analyses, miR-21 and miR-126 significantly impacted CSS in our cohort. Moreover, applying the miR-based risk classifier significantly stratified ccRCC^IVC^ according to CSS.

**Conclusions:** In our retrospective analysis, we successfully validated the miR-based risk classifier within an independent ccRCC^IVC^ cohort.

## 1. Introduction

In about 10% of all cases, clear cell renal cell carcinomas (ccRCC) extend into the inferior vena cava (ccRCC^IVC^) [1–3]. While constituting a high-risk setting in general, there still is substantial clinical heterogeneity within the ccRCC^IVC^ subgroup – with reported 5-year survival rates ranging from 37% to 65% for non-metastasized patients treated with nephrectomy in combination with tumor thrombectomy [4–9]. Regarding this discrepancy, biomarkers are urgently needed to identify patients with a specifically high risk of cancer relapse [10,11]. Potentially, ccRCC^IVC^ patients may also benefit from adjuvant systemic therapy and an intensified follow-up.

MicroRNAs (miRs) as biomarker candidates are post-transcriptional regulators of gene expression in various cancer entities [12–14]. Regarding ccRCC, several studies demonstrated the prognostic impact of miR expression levels in tumor tissue [15–17]. Previously, Vergho et al. established a combined risk classifier for patients with ccRCC^IVC^ receiving nephrectomy and thrombectomy in curative intention [10]. Based on miR-21, miR-126 and miR-221 expression in tumor tissue, the risk classifier significantly stratified patients regarding cancer-specific survival (CSS) in a single-center cohort (*n* = 37) – with a 5-year CSS of 78% vs. 18% in the favorable compared to the unfavorable subgroup [10].

To further assess the miR-based risk classifier as a prognostic tool in ccRCC^IVC^ patients, we retrospectively evaluated it within an independent cohort (*n* = 56) from the Department of Urology, University of Regensburg (Regensburg, Germany). Cut-off values for miR expression levels as well as internal classifier weights were transferred from the previous pilot study [10], in order to test its transferability to independent study cohorts. Figure 1 illustrates the course of our study.

**Figure 1.**
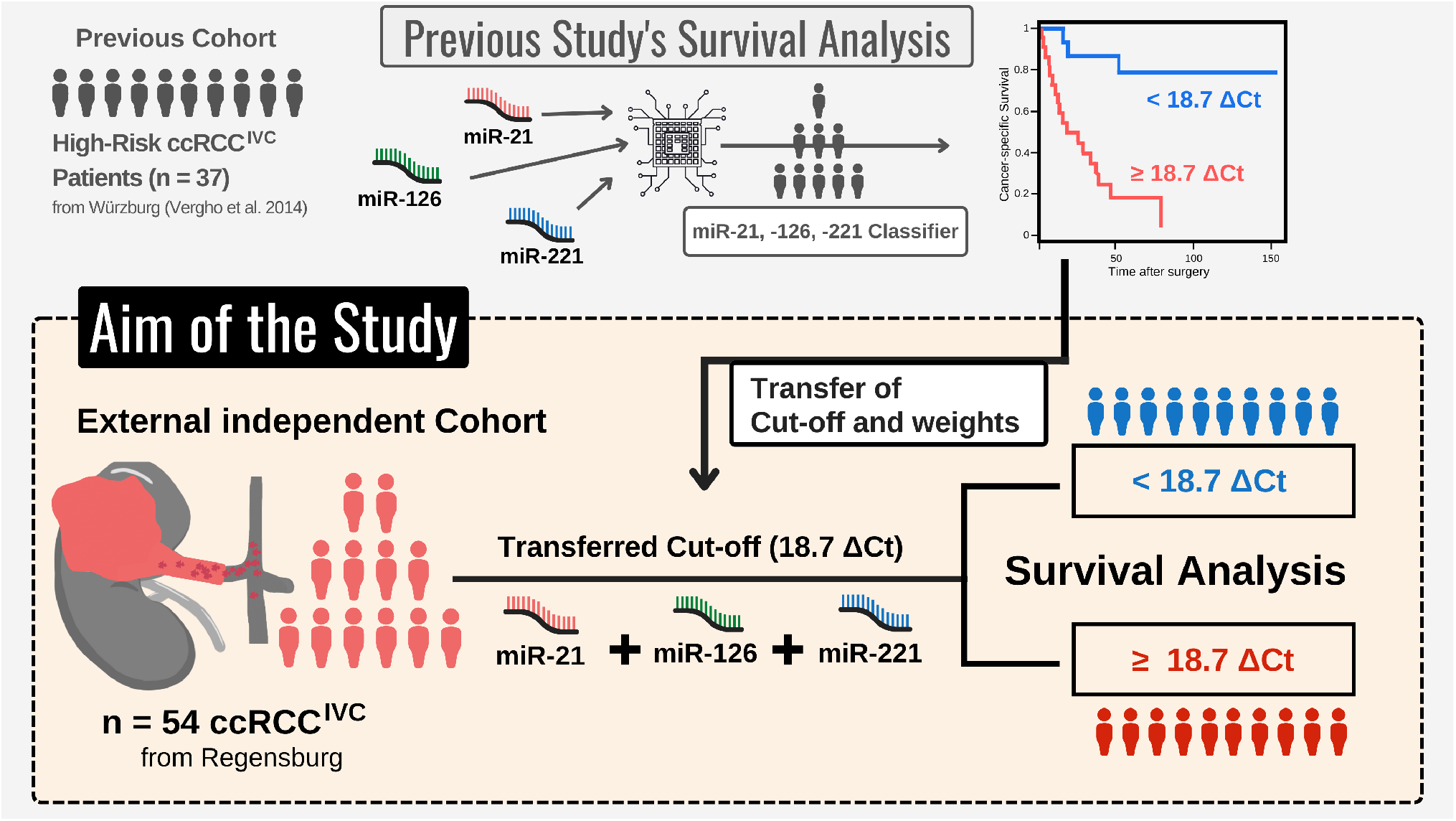
Course of the study – using a microRNA (miR)-based risk classifier established previously [10], we examined the prognostic impact of miR-21, miR-126, and miR-221 expression in an independent cohort of clear cell renal cell carcinoma samples with infiltration of the inferior vena cava (ccRCC^IVC^; *n* = 54). To assess the transferability of the miR-based risk classifier, cut-off value and weights were identical to the previous study.

## 2. Materials and Methods

### 2.1. Tumor Tissue Samples and Patients

Paraffin-embedded primary ccRCC^IVC^ tumor-samples of 56 subjects who underwent radical surgery were aggregated by the Department of Urology, University of Regensburg, Germany (1997– 2006). A uropathologist selected sample-regions with > 90% cancerous tissue. Follow-up data were collected by the Department of Urology, University of Regensburg (Regensburg, Germany). The study was approved by the local Ethics Committee (Regensburg: Nr. 08/108). Detailed characteristics of the study cohort are summarized in Table 1.

**Table 1.** Clinical and pathological patient characteristics (*n* = 56). Detailed follow-up information was available for 54 patients.

| Characteristics |  |
| --- | --- |
| **Median Follow-up** | 94 (1 – 190) months |
| **Median Age** | 67 (41 – 89) years |
| **Sex** |  |
| female | 22 (39.3%) |
| male | 34 (60.7%) |
| **Tumor Stage: pT3b** | 56 (100%) |
| **Fuhrman Grade** |  |
| G2 | 41 (73.2%) |
| G3 | 15 (26.8%) |
| **Nodal Status** |  |
| N0 | 45 (80.4%) |
| N+ | 11 (19.6%) |
| **Distant Metastasis during Follow-up** |  |
| M0 | 35 (62.5%) |
| M1 | 21 (37.5%) |
| **Median Tumor Size** | 70 (18 – 225) mm |
| **Overall survival** |  |
| yes | 27 (48.2%) |
| no | 29 (51.8%) |
| **Cancer-related death** |  |
| yes | 13 (23.2%) |
| no | 43 (76.8%) |

### 2.2. RNA Extraction and qRT-PCR

Using the RecoverAll™ Total Nucleic Acid Isolation Kit (Thermo Fisher Scientific, Waltham, MA), total RNA from paraffin-embedded samples was isolated according to the manufacturer’s instructions. RNA concentration and 260/280 ratio were analyzed by Spark^®^ 10M (TECAN, Männedorf, Switzerland). cDNA was synthesized from total RNA with stem-loop reverse transcription primers (TaqMan microRNA Assay protocol, Applied Biosystems, Birchwood, UK). TaqMan microRNA Assay kit was used to quantify miR expression according to manufacturer’s protocols. Samples showing a standard deviation > 0.5 were excluded (all reactions performed in triplicates). Small nuclear RNA (RNU6B) expression was used for normalization of miRs relative expression values. Samples with expression levels of RNU6B > 30 Ct were excluded from further analyses. Relative miR expressions were calculated using the ΔCt-method (ΔCt sample = Ct miR of interest − Ct RNU6B). To calculate fold changes in miR expression between samples, we used the 2ΔΔCt method (in this study, referred to as the ΔΔCt method).

### 2.3 Statistics and computational analysis

A Jupyter Notebook environment (version 6.3.0) was used to perform all statistical analyses using Python version 3.8.8, LifeLines version 0.27.0 [18], Pandas version 1.2.4 [19], Matplotlib 3.3.4 [20], Scipy version 1.6.2 [21].

To analyze differences between miR expression levels, we used Student’s t-test for normally distributed data with similar variance – otherwise, Mann-Whitney-U-Test was applied. Data distribution and variance were assessed via Shapiro Wilk and Levene test, respectively. A significance level of 0.05 was applied.

#### 2.3.1 Validation of microRNA-based Risk Classifier

To evaluate the validity of the miR-based risk classifier – (4.592 × ΔCt miR-21) + (−3.892 × ΔCt miR-126) + (−1.938 × ΔCt miR-221) – weights and cut-off values (≥ 18.7 ΔCt = “unfavorable subgroup”, < 18.7 ΔCt = “favorable subgroup”) were transferred from the pilot study [10] and applied in order to stratify the ccRCC^IVC^ study cohort from Regensburg and perform Kaplan-Meier analyses. Within the risk classifier formula, a negative factor indicates that higher expression levels correlate with longer survival, while a positive factor correlates with shorter survival. For further analysis, we transferred cut-off values for miR-21 (8.17 ΔCt), miR-126 (3.57 ΔCt) and miR-221 expression (1.84 ΔCt) to evaluate their predictive potential in the new cohort using the Kaplan-Meier survival analysis.

#### 2.3.2 microRNA-based Risk Classifier Calculation

Referring to the pilot study by Vergho et al. [22], the risk classifier was calculated as follows:

1. Performing uni- and multivariate Cox regression analysis, Vergho et al. evaluated the impact of clinicopathological parameters and various miRs on CSS.
2. To select the best fitting Cox model, the relative goodness-of-fit was measured based on the Akaike information criterion. The combination of miR-21, -126 and -221 displayed the best prediction properties.
3. Finally, the miR-based risk classifier was calculated based on the publication by Lossos et al. [23]. Hereby, a factor attained from the Cox model’s z-score was identified for miR-21, -126 and -221. In the next step, relative expression levels (ΔCt) of miRs were multiplied by these factors (weights) using the formula (4.592 × ΔCt miR-21) + (−3.892 × ΔCt miR-126) + (−1.938 × ΔCt miR-221). Risk score cut-off (18.7 ΔCt) was determined by ROC.

## 3. Results

Table 1 summarizes the basic clinical and pathological characteristics of our study cohort. Detailed follow-up information was available for 54 of 56 patients suffering from ccRCC^IVC^ (96.4%).

### 3.1. Association of miR-21, -126 and -221 Expression with Clinicopathological Characteristics

To investigate the impact of miR-21, -126 and -221 within our ccRCC^IVC^ cohort, we associated expression levels of miR-21, miR-126 and miR-221 to relevant clinical parameters. Figure 2 illustrates the results.

**Figure 2.**
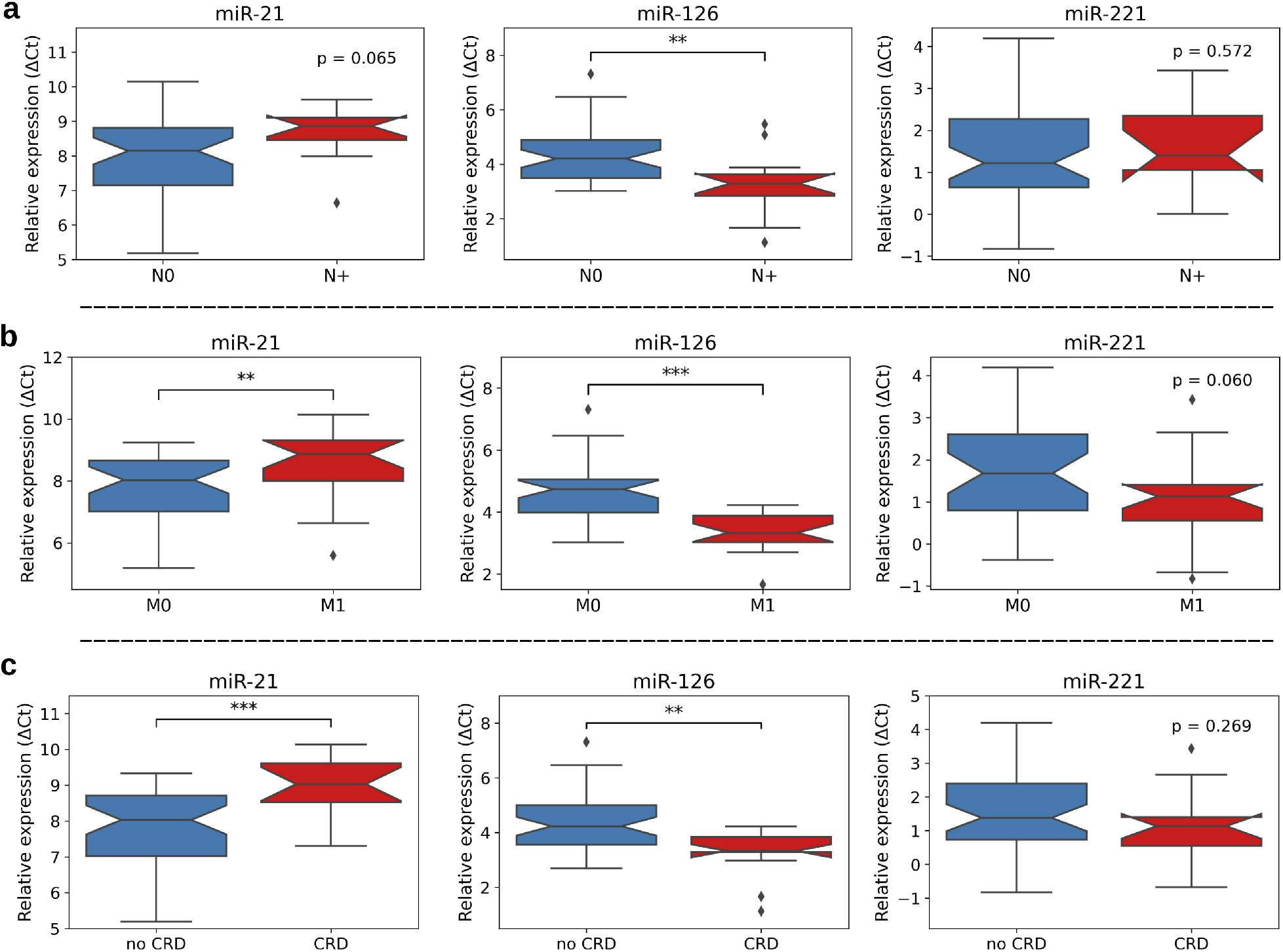
miR-21, -126 and -221 expression levels depending on lymphonodal status (a), distant metastases (b) and cancer-related death (CRD, c). Significant changes between subgroups were calculated using unpaired Student’s t test (CRD: miR-221; nodal status: miR-21, -221; distant metastases: miR-21, -126) or Mann-Whitney-U test (CRD: miR-21, -126; nodal status: miR-126; distant metastases: miR-221). *p* < 0.05 *; *p* < 0.01 **; *p* < 0.001 ***.

At time of surgery, 11 of 56 ccRCC^IVC^ patients (19.6%) were diagnosed with nodal metastasis. In cases with positive nodal status, a trend towards up-regulation of miR-21 (*p* = 0.065) and a significant down-regulation of miR-126 (*p* < 0.01) were observed. For miR-221, there was no statistically significant association to nodal status.

During the follow-up period, distant metastasis emerged in 21 of 56 ccRCC^IVC^ patients (37.5%). As shown in Figure 2b, we observed a significant up-regulation of miR-21 (*p* < 0.01) and down-regulation of miR-126 (*p* < 0.001) as well as a trend towards downregulation for miR-221 (*p* = 0.06) in ccRCC^IVC^ samples of patients with metastasized disease.

Of 56 patients with ccRCC^IVC^, 13 (23.2%) died during the follow-up period due to cancer (cancer-related death, CRD). Regarding miR expression levels, we found a significant up-regulation of miR-21 (*p* < 0.001) and a down-regulation of miR-126 (*p* < 0.01) in CRD cases. Instead, miR-221 expression did not show a statistically significant association to CRD in this analysis (*p* = 0.27).

### 3.2. Cox regression Analysis

Next, we performed a univariate Cox regression analysis to further assess the prognostic potential of miR-21, -126, and -221 expression levels as predictors of CRD. Detailed follow-up information was available for 54 of 56 cases with a median of 94 months.

As summarized in Table 2a, miR-21 and miR-126 significantly predicted the occurrence of CRD in our study cohort (*p* = 0.003, Hazard Ratio (HR) 3.79 for miR-21, *p* = 0.00003, HR 0.19 for miR-126). In contrast, miR-221 expression in tumor tissue did not display significant prognostic potential (*p* = 0.22). Regarding further clinical parameters, significant results were also observed for nodal involvement, metastatic status and Fuhrman grade.

**Table 2.** (a) Univariate Cox regression of ccRCC^IVC^ patients for miR expression levels and clinicopathological parameters, (b) multivariate Cox regression for miR expression levels as well as Nodal Status and Fuhrman Grade. 95% Confidence intervals (CI) shown for Hazard ratios (HR). *p* values were computed using the chi-squared test. *p* < 0.05 *; *p* < 0.01 **; *p* < 0.001 ***.

| Parameters | Cancer-related death |  |  |  |
| --- | --- | --- | --- | --- |
|  | Univariate analysis |  | (b) Multivariate analysis |  |
| | HR (95% CI) | $p$ value | HR (95% CI) | $p$ value |
| miR-21 | 3.79<br>(1.55 – 9.26) | **0.003\*\*** | 4.94<br>(1.29 – 18.98) | **0.02\*** |
| miR-126 | 0.19<br>(0.09 – 0.42) | **0.00003\*\*\*** | 0.27<br>(0.097 – 0.75) | **0.01\*** |
| miR-221 | 0.74<br>(0.46 – 1.19) | 0.22 | 0.64<br>(0.37 – 1.12) | 0.12 |
| Age | 0.98<br>(0.92 – 1.03) | 0.42 |  |  |
| Sex | 2.10<br>(0.58 – 7.65) | 0.26 |  |  |
| Tumor size | 1.01<br>(1.00 – 1.03) | 0.07 |  |  |
| Fuhrman Grade | 3.79<br>(1.27 – 11.33) | **0.02\*** | 3.28<br>(0.70 – 15.32) | 0.13 |
| Nodal status | 6.70<br>(2.09 – 21.47) | **0.001\*\*** | 1.34<br>(0.29 – 6.12) | 0.71 |
| Distant metastasis | ∞ | NA | | |

Abbreviation: CI, confidence interval; HR, hazard ratio; ∞ / NA: In case of distant metastasis as predictor of CRD, the coefficient was not estimable (positively infinite). HR and $p$ values were therefore not shown.

Additionally, as shown in Table 2b, we performed a multivariate Cox regression to investigate if the miR candidates remain acting as relevant predictors of CRD within our ccRCC^IVC^ cohort in attendance of nodal involvement and Fuhrman grade. Also under this conditions miR-21 and miR-126 significantly predicted the occurrence of CRD (*p* = 0.02, HR 4.94 for miR-21, *p* = 0.01, HR 0.27 for miR-126). MiR-221 expression again did not meet statistical significance as predictor of CRD (*p* = 0.12) (Table 2 and Figure 3d). No significant results were observed for the clinical parameters nodal involvement (*p* = 0.71) and Fuhrman grade (*p* = 0.13).

**Figure 3.**
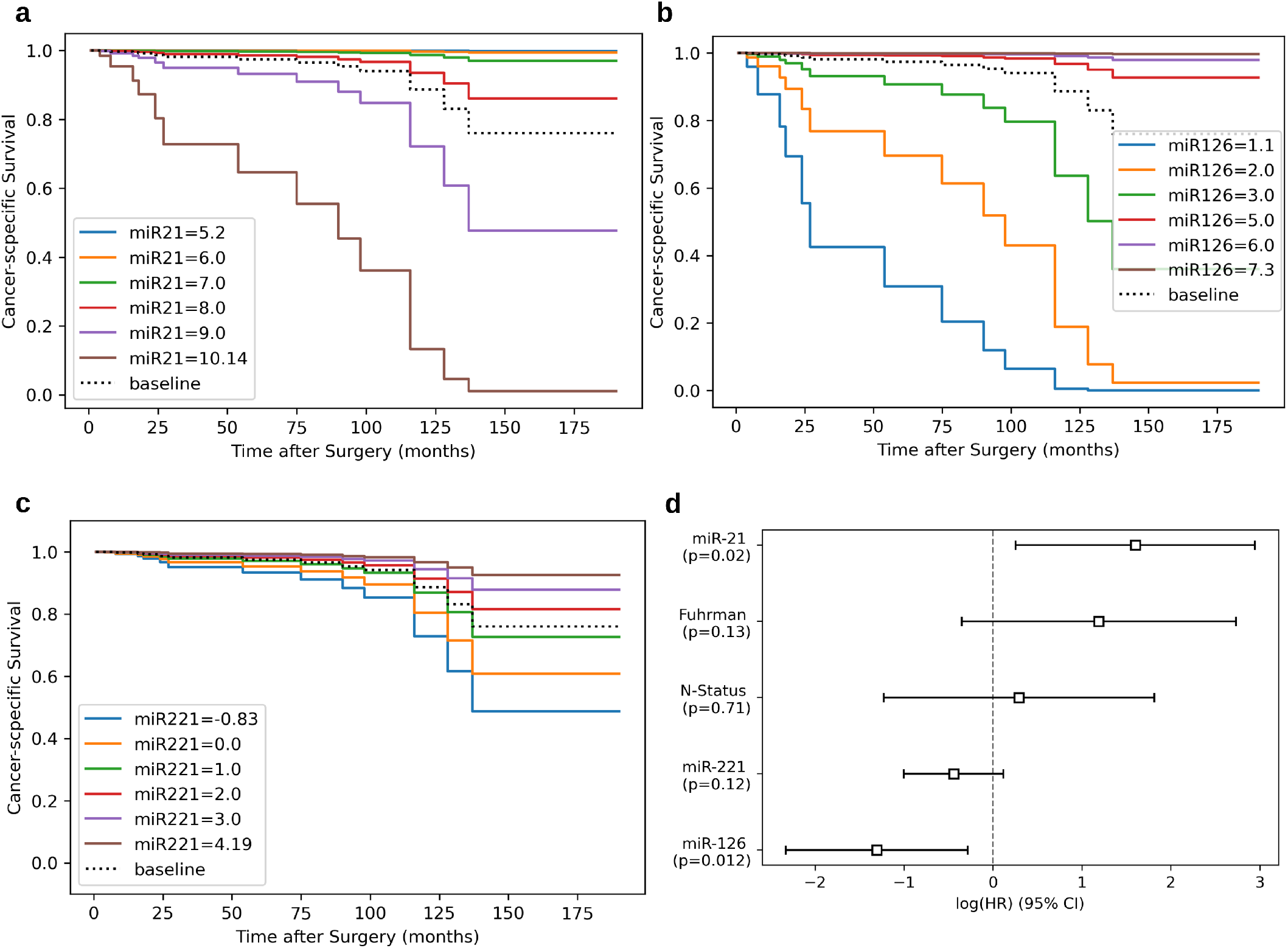
(a–c) Survival curves for varying miR expression levels of miR-21, -126, -221 (based on fitted multivariate Cox regression model), illustrating partial effects of single miRNAs on cancer-specific survival for present study cohort (*n* = 54). The baseline represents median relative expression for each miRNA (miR-21 = 8.34 ΔCt, miR-126 = 4.0 ΔCt, miR-221 = 1.3 ΔCt). Blue line represents minimum and brown line the maximum relative expression of each miRNA in present study cohort. Other expression levels were chosen randomly (1.0 ΔCt steps). (d) Forest plot representing log Hazard Ratios (HR) from multivariate Cox regression of miR-21, miR-126, miR-221, Fuhrman Grade (Fuhrman) and nodal status (N-Status) for cancer-related death (CRD). *p* values were computed using the chi-squared test.

To further understand and illustrate the prognostic impact of single miRNAs on CSS (based on previously fitted multivariate Cox regression model), we plotted survival curves according to isolated miR expression levels (Figure 3a-c). For miR-21, higher relative expression levels were associated with lower CSS (Figure 3a). In contrast, higher expression levels of miR-126 as well as miR-221 were associated with higher CSS (Figure 3b, c).

### 3.3. Kaplan Meier Analyses for single miR expression and the Risk Classifier

To investigate the prognostic transferability of single miRNAs, survival analyses using the cutoffs from Vergho et al. [10] were performed. Both, miR-21 and miR-126 showed a strong predictive significance in the Kaplan-Meier survival analysis (Fig. 4a, b). However, differences regarding CSS of miR-221 high vs. low expressing tumor specimens (Fig. 4c) did not reach statistical significance (*p* = 0.25).

**Figure 4.**
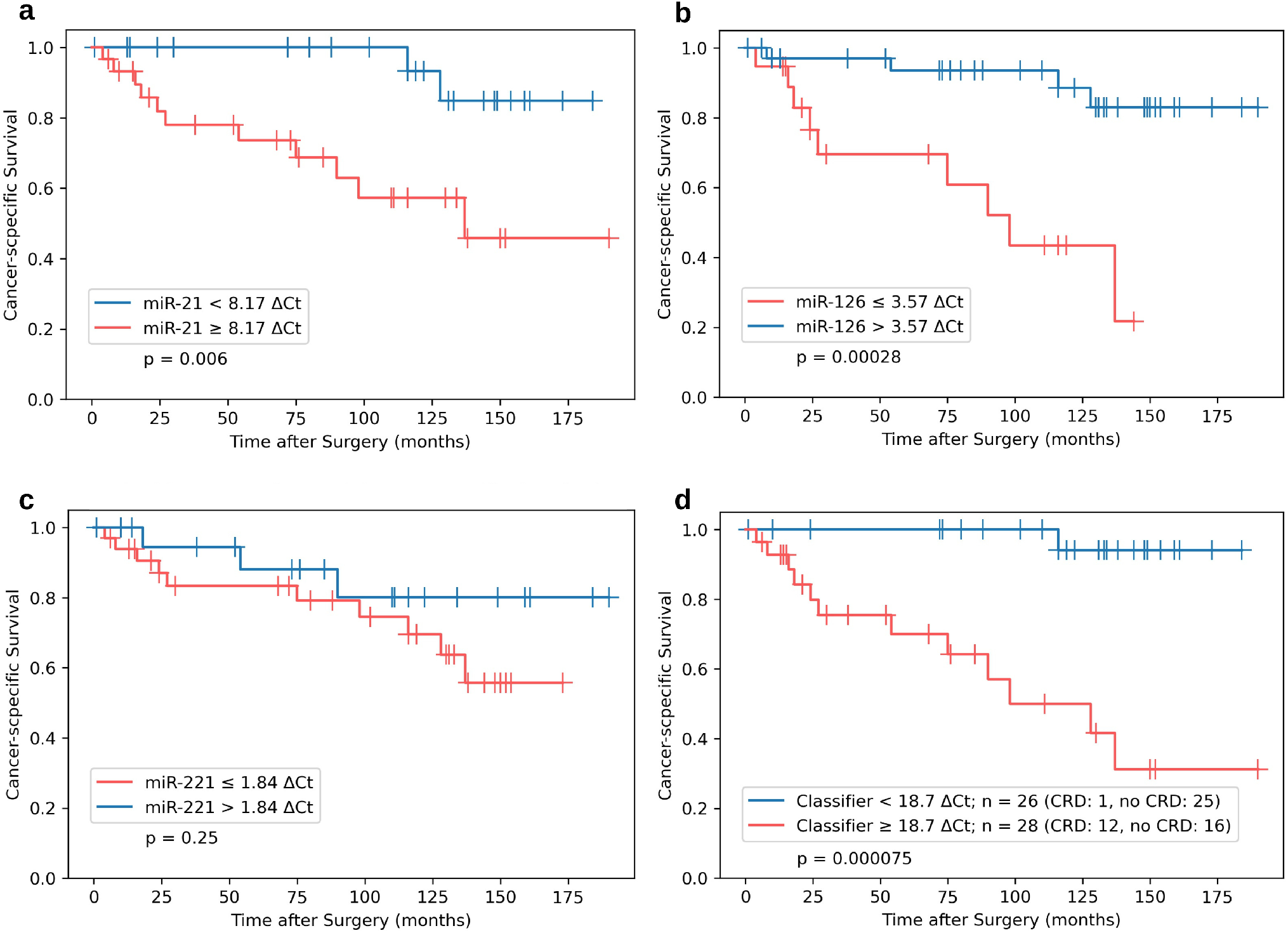
Kaplan Meier survival analysis for CSS for external independent ccRCC^IVC^ (*n* = 54) cohort from Regensburg stratified by miR-21 (a), miR-126 (b) and miR-221 (c) expression levels. (d) Combined miR-based risk classifier (miR-21, -126, -221) using identical cut-offs and weights from a previous publication [10]. *p* values from log-rank tests are shown within each plot.

To validate the predictive potential of the risk classifier (Fig. 4d), the established cut-off level of 18.7 ΔCt (≥ 18.7 ΔCt = unfavorable subgroup, < 18.7 ΔCt = favorable subgroup) was used to stratify the validation cohort consisting of *n* = 54 ccRCC^IVC^ tissue samples. Out of 13 CRD cases, the classifier correctly identified 12 patients who suffered from CRD as members of the unfavorable subgroup (92.3%; ≥ 18.7 ΔCt). Overall, a sensitivity of 92.3% (CI 95%: 62.1% – 99.6%) and a specificity of 61.0% (CI 95%: 44.5% – 75.4%) were reached. Difference in 5-years and 10-years CSS was 100% vs. 70% and 94% vs. 31% between the favorable and the unfavorable subgroup, respectively.

## 4. Discussion

ccRCCs infiltrating the inferior vena cava represent a clinically relevant high-risk subgroup. Still, there is substantial clinical heterogeneity within this distinct subgroup – and biomarkers are needed to assess the individual risk of progression. In general, adjuvant therapy with tyrosine kinase inhibitors (TKI) or immune checkpoint blockers could be a promising therapeutic option after nephrectomy – especially for patients suffering from high-risk RCC. However, European kidney cancer guidelines currently do not contain strong recommendations towards adjuvant therapies due to the mixed outcome in clinical trials [24]. For the TKI sunitinib, one trial displayed improved disease-free survival (DFS) for patients – while showing no significant differences in overall survival (OS) [25]. Additionally, another phase 3 trial did not detect significant survival effects for adjuvant sunitinib or sorafenib in nonmetastatic high-risk renal cell carcinoma [26]. Due to the sobering TKI results, research efforts were mainly shifted towards immune checkpoint blockers. For the PD-1 (Programmed Cell Death Protein 1) inhibitor pembrolizumab, KEYNOTE-564 trial detected improved progression-free survival (PFS) in an adjuvant setting after nephrectomy [27].

### 4.1. Evaluating a miR-based Risk Classifier for RCC with Infiltration of the Vena Cava

To estimate the individual risk of patients suffering from ccRCC^IVC^, members of our research group have established a risk classifier based on the tissue expression of miR-21, miR-126, and miR-221 [10]. Former cohort contained tumor tissue of *n* = 37 patients undergoing surgery at the University Hospital of Würzburg, Germany. In this study, we externally validated the prognostic potential of the miR-based risk classifier. Therefore, we examined an independent cohort of ccRCC^IVC^ from the University of Regensburg, Germany (*n* = 56). To test the transferability and usability of the classifier within an external tissue cohort, we applied identical cut-off values and weights as in the previous pilot study.

Regarding clinicopathological characteristics of our study cohort, low miR-126 expression was significantly associated with a positive lymphonodal status at time of surgery. Moreover, occurrence of metastases during follow-up was significantly associated with higher miR-21 and lower miR-126 levels. Tumor tissue from patients suffering from CRD was also characterized by a significant upregulation of miR-21 and a downregulation of miR-126. Within univariate cox regression, miR-21 and miR-126 showed prognostic significance regarding cancer-specific survival (CSS). Lower levels of miR-221 in tumor tissue and its association with CRD did not reach statistical significance. Beyond miR expression, Fuhrman grade, lymphonodal status and occurrence of metastasis emerged as prognostically relevant. Next, we added a multivariate Cox regression for the three miR candidates. Again, this study identified miR-21 and miR-126 expression to significantly impact CRD. Kaplan Meier analyses based on cut-off values determined previously by Vergho et al. [10] confirmed the significant influence of miR-21 as well as miR-126 expression levels on CSS. Finally, applying the miR-based classifier using identical cut-off values and weights split patients in two groups.

Of note, the classifier nearly stratified the study cohort in two halves – with *n* = 26 patients belonging to the favorable and *n* = 28 patients belonging to the unfavorable subgroup. Regarding the substantial difference in CSS between both groups, adjuvant therapies appear promising especially for the unfavorable subgroup of ccRCC^IVC^ patients.

### 4.2. Functional Roles of miR-21, miR-126, and miR-221 in Cancer

After confirming the prognostic potential of the miR classifier using the validation cohort from Regensburg, we were interested in previously reported functions of these miRs in RCC and other malignancies. For miR-21, several researchers demonstrated oncogenic effects in various cancers, including RCC [28,29]. Among the prominent miR-21 target genes are key players of apoptosis induction like PDCD4 (Programmed Cell Death 4) [30] and genes like PTEN (Phosphatase and Tensin Homolog) [28]. Latter is an established tumor suppressor gene best known for regulating PI3K/Akt signaling. In contrast to miR-21, miR-126 acts as a tumor suppressor in tumor tissue, e. g. by targeting ROCK1 (Rho Associated Coiled-Coil Containing Protein Kinase 1) [31] and VEGFA (Vascular Endothelial Growth Factor A) [32]. For miR-221, oncogenic versus protective functions appear to depend on the underlying cancer entity, as researchers demonstrated both roles [33–35]. For RCC, a down-regulation of miR-221 appears well in line with previous publications. Specifically, miR-221 is reported to regulate KDR (Kinase Insert Domain Receptor) – also known as VEGFR2 (Vascular Endothelial Growth Factor Receptor 2) – in ccRCC [36] and prostate cancer [37], thereby regulating the sensitivity towards sunitinib. In summary, among diverse tumorigenic functions of these miRs, all three candidates prominently influence angiogenesis-related pathways (so-called AngiomiRs) [38,39]. Given that not all ccRCCs depend on angiogenic signaling to the same degree [40], it is tempting to assume that the unfavorable subgroup identified by our risk classifier could benefit from adjuvant anti-angiogenic therapy.

### 4.3. Limitations and Future Directions

Our study has several limitations. Leaving aside the definite RCC subgroup investigated here, sample size of our study is relatively small. Moreover, we purposely did not adjust cut-off values and individual miR weights determined previously in order to check the transferability of the risk classifier to external tissue cohorts. More research – ideally in a prospective setting – could further validate the risk classifier in a clinical setup and elucidate whether sub-classification of ccRCC^IVC^ is able to identify patients most susceptible towards adjuvant therapy.

## 5. Conclusions

While RCC extending into the inferior vena cava represents a high-risk setting, there is still substantial clinical heterogeneity within this patient subgroup. Previously, Vergho et al. established a miR-based risk classifier – containing miR-21, miR-126 and miR-221 expression – which significantly predicted CSS for patients from this subgroup. To validate this classifier, we examined its impact on an external and independent patient cohort. Using identical cut-off values for single miRs and identical weights within the classifier, we confirmed a highly significant risk stratification within the new cohort. Patients with an unfavorable constellation according to the miR-based classifier could especially benefit from adjuvant therapy and continuous follow-up examinations.

## Data Availability

All data produced in the present study are available upon reasonable request to the authors.

## Author Contributions

Conceptualization, M.Ko., M.Kr., B.K., D.V. and C.K.; methodology, M.Kr., B.K., D.V. and C.K.; software, M.Ko., S.K. and B.K.; validation, M.Kr., M.B., J.B., H.K., D.V., B.K. and C.K.; formal analysis, M.Ko., S.K. and C.K.; investigation, M.Ko., M.Kr., W.O., J.B., S.K., B.K. and C.K; resources, J.B. and B.K.; data curation, M.Ko., B.K. and C.K.; writing—M.Kr., M.Ko., J.B., B.K. and C.K.; writing—review and editing, M.Ko., M.Kr., M.B., H.K., R.B., W.O., J.B., S.K., D.V., B.K. and C.K.; visualization, M.Ko. and M.Kr.; supervision, M.B., H.K., R.B., W.O., D.V., B.K. and C.K.; project administration, M.Kr., B.K. and C.K.; funding acquisition, M.B. and H.K.

## Funding

Markus Krebs was funded by a personal grant from Else-Kröner-Foundation (Else Kröner Integrative Clinician Scientist College for Translational Immunology, University Hospital Würzburg, Germany).

## Institutional Review Board Statement

All procedures performed in studies involving human participants comply with the ethical standards laid down in the latest declaration of Helsinki of the World Medical Association. The study was approved by the local Ethics Committees (Regensburg: Nr. 08/108, Würzburg: Nr. 136/08). All involved patients gave their written and informed consent for the publication of their anonymized cases.

## Informed Consent Statement

Informed consent was obtained from all subjects involved in the study.

## Data Availability Statement

All data produced in the present study are available upon reasonable request to the authors.

## Conflicts of Interest

The authors declare no conflict of interest.

## Notes

### Competing Interest Statement

The authors have declared no competing interest.

### Funding Statement

Markus Krebs was funded by a personal grant from Else-Kroener-Foundation (Else Kroener Integrative Clinician Scientist College for Translational Immunology, University Hospital Wuerzburg, Germany).

### Author Declarations

All procedures performed in studies involving human participants were in accordance with the ethical standards of the institutional and/or national research committee and with the 1964 Helsinki declaration and its later amendments or comparable ethical standards. The study was approved by the local Ethics Committees (Regensburg: Nr. 08/108, Wuerzburg: Nr. 136/08).

### Summary of Updates

The Material and Methods were updated to clarify the calculation of the microRNA-based Risk Classifier. A multivariate Cox Regression Analysis was added to the Results section. Description of Figure 3 was extended.

